# An Assessment of Transparency and Reproducibility-related Research Practices in Otolaryngology

**DOI:** 10.1101/19002238

**Authors:** Austin L. Johnson, Trevor Torgerson, Mason Skinner, Tom Hamilton, Daniel Tritz, Matt Vassar

**Affiliations:** Oklahoma State University Center for Health Sciences, Tulsa, Oklahoma; Oklahoma State University Medical Center, Department of Otolaryngology, Tulsa, Oklahoma

**Author notes:** Corresponding Author: Austin L. Johnson.; 1111 W 17th St. Tulsa, OK 74137. Data Availability Statement: All protocols, materials, and raw data are available online (https://osf.io/x24n3/). Funding and Conflict of Interest: This study was funded through the 2019 Presidential Research Fellowship Mentor–Mentee Program at Oklahoma State University Center for Health Sciences. We declare no conflicts of interest. Author Approval: All authors have seen and approve of this manuscript.

**Keywords:** Reproducibility, Replication, Otolaryngology, Open Science, Data Sharing, Protocol, Open Access

## Abstract

**Introduction:** Clinical research serves as the foundation for evidence-based patient care, and reproducibility of results is consequently critical. We sought to assess the transparency and reproducibility of research studies in otolaryngology by evaluating a random sample of publications in otolaryngology journals between 2014 and 2018.

**Methods:** We used the National Library of Medicine catalog to identify otolaryngology journals that met the inclusion criteria (available in the English language and indexed in MEDLINE). From these journals, we extracted a random sample of 300 publications using a PubMed search for records published between January 1, 2014, and December 31, 2018. Specific indicators of reproducible and transparent research practices were evaluated in a blinded, independent, and duplicate manner using a pilot-tested Google form.

**Results:** Our initial search returned 26,498 records, from which 300 were randomly selected for analysis. Of these 300 records, 286 met inclusion criteria and 14 did not. Among the empirical studies, 2% (95% CI, 0.4%-3.5%) of publications indicated that raw data were available, 0.6% (95% CI, 0.3%-1.6%) reported an analysis script, 5.3% (95% CI, 2.7%-7.8%) were linked to an accessible research protocol, and 3.9% (95% CI, 1.7%-6.1%) were preregistered. None of the publications had a clear statement claiming to replicate, or to be a replication of, another study.

**Conclusions:** Inadequate reproducibility practices exist in otolaryngology. Nearly all studies in our analysis lacked a data or material availability statement, did not link to an accessible protocol, and were not preregistered. Most studies were not available as open access. Taking steps to improve reproducibility would likely also improve patient care.

## INTRODUCTION

Clinical research serves as the foundation for evidence-based patient care. Given the important role of research in establishing standards of care, reproducible research is critical. However, some authors have expressed concerns that many published research findings may be false or irreproducible.^1–3^ In the field of otolaryngology, for example, a study published in *JAMA Otolaryngology* that investigated the use of dexamethasone for complications post thyroid surgery was recently retracted, partly because of incorrect statistical results that did not support the authors’ conclusions.^4,5^ Prior to retraction, the journal requested an investigation into the legitimacy of the authors’ findings. The investigation was never completed because the authors failed to provide the raw data and the original protocol.^4^ Access to raw data sets and protocols—2 components of transparent and reproducible science—is of paramount importance in reducing and correcting false research findings

Overall, only 10% to 25% of biomedical research findings are estimated to be reproducible,^6–9^ and scientific research is thus experiencing a “reproducibility crisis.”^6^ Independent verification of results (ie, reproducibility) is key to the scientific process and aids in enhancing the credibility, utility, and reliability of published literature.^10^ A survey conducted by the editorial staff of the journal *Nature* found that more than 70% of the 1500 scientists surveyed were unable to replicate the same outcomes from another scientist’s experiment, and half of these scientists reported failures replicating their own experiments.^11^ A project dedicated to reproducing key findings in cancer biology experiments was recently abandoned after 32 of 50 replication attempts failed, partly because scientists were unable to obtain sufficient detail to reproduce the original studies.^12^ These results suggest significant room exists for improvement with regard to reproducible research.

Irreproducible studies often stem from incomplete reporting of methodology or failing to provide access to materials, protocols, detailed methods, or raw datasets.^13,14^ A study comprising 198 publications related to social science reported that only 16% of publications provided access to materials, a few (8%) provided access to raw datasets, none provided access to protocols, and only 3% provided access to analysis scripts.^15^ Likewise, a study of biomedical journal publications found that only 1 study provided a full protocol, none made the raw data available, and only 4 were replication studies.^16^ Because experimental methodologies are often complex, critically assessing the accuracy of scientific claims and conclusions drawn from them is difficult without access to all relevant study materials.^17^ We should question why published scientific research has taken this direction.

The extent of reproducible practices in the field of otolaryngology remains unknown. To date, no studies have explored current practices of the otolaryngology research community with regard to reproducible research practices being included in published results. Here, we evaluated a random sample of the otolaryngology literature using indicators of reproducibility and transparency established by Hardwicke et al.^15^ Our goals were to elucidate areas in need of improvement and to establish baseline data for subsequent investigations of the otolaryngology literature.

## METHODS

We used the protocol of Hardwicke et al,^15^ with modifications explained below, for the present investigation. Because our study did not include human participants, it was not subject to institutional review board oversight per US Department of Health and Human Services’ Code of Federal Regulation 45 CFR 46.102(d) and (f).^18^ This study is reported according to the guidance developed by Murad and Wang^19^ for meta-research studies and, when relevant, the Preferred Reporting Items for Systematic Reviews and Meta-Analyses (PRISMA).^20^ Our protocol, raw data, and other pertinent materials have been deposited on the Open Science Framework (https://osf.io/x24n3/), which has introduced new infrastructure supporting research transparency. Our primary endpoint was an evaluation of indicators of reproducible and transparent research practices within otolaryngology.

### Journal Selection

On May 29, 2019, one of us (DT) conducted a detailed search using the National Library of Medicine catalog for all journals, using the subject term tag otolaryngology[ST]. For inclusion, each journal had to be available in the English language and indexed in MEDLINE. Using this list of journals, an advanced search on PubMed (which catalogues the entire MEDLINE database) was conducted by DT using the electronic or, if unavailable, the linking ISSN numbers of each journal on May 31, 2019. We limited our search to publications between January 1, 2014, and December 31, 2018. We used these search results to obtain a random sample of 300 publications by applying Excel’s random number function.

### Study Selection

To incorporate a wide range of publications, we did not specify particular study designs for inclusion. Rather, we included all studies with empirical data for analysis.

### Training

Prior to data extraction, 2 of us (AJ, TT) underwent rigorous training to ensure reliability between investigators. The training included an in-person session to review the study design, protocol, extraction form, and location of the information within publications. During training, the 2 investigators extracted data from 2 publications independently; after which, results were compared, and disagreements were reconciled. The investigators then extracted data from 10 additional studies, using the same blinded and double data extraction process. Consensus was achieved by discussion. This training session was recorded and posted online for reference.

### Data Extraction

Following training, AJ and TT extracted data from the 300 included publications in a duplicate and blinded fashion. Data extraction was performed between June 3 and 13, 2019, using a pilot-tested Google form. After data extraction was completed, a final consensus meeting was held by the pair to resolve disagreements. A third investigator (DT) was available for adjudication but was not ultimately needed. During extraction, publications were separated into 2 categories: publications with no empirical data (editorials, commentaries [without reanalysis], simulations, news, reviews, and poems) and publications with empirical data (clinical trial, cohort, case series, case reports, case-control, secondary analysis, chart review, commentary [with data analysis], and cross-sectional). For all empirical and nonempirical publications that we could access, we extracted the funding source, conflict of interest declarations, and impact factor (2016 or 2017 and 5-year impact factors). The variables extracted for empirical publications varied by their study design. For example, we did not evaluate case studies for the presence of a protocol because such studies are unlikely to have a prespecified protocol because they present interesting patient cases as they occur in clinical work.^21^ The data extraction protocol for each study design is outlined in Table 1.

**Table 1:**
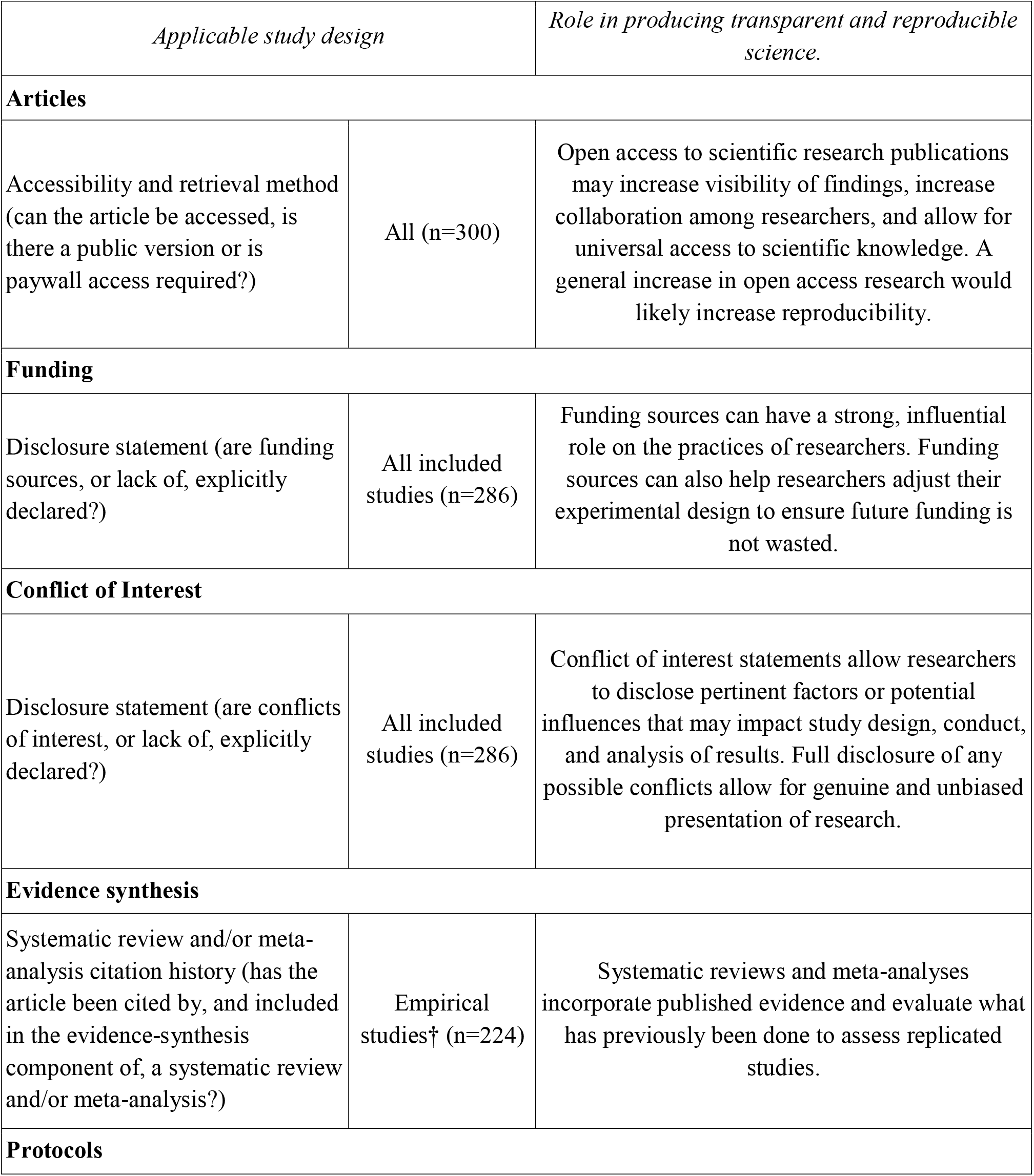
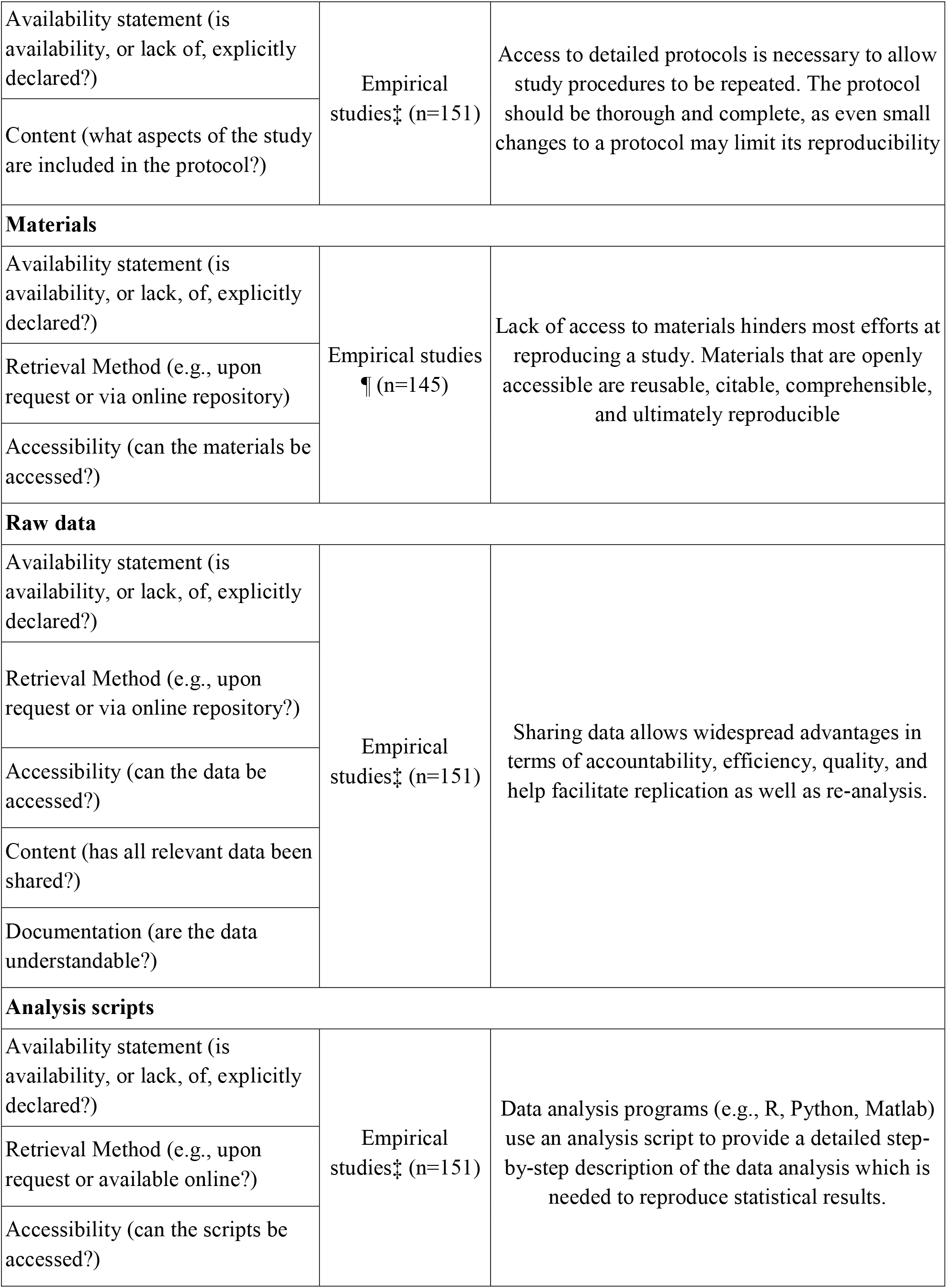
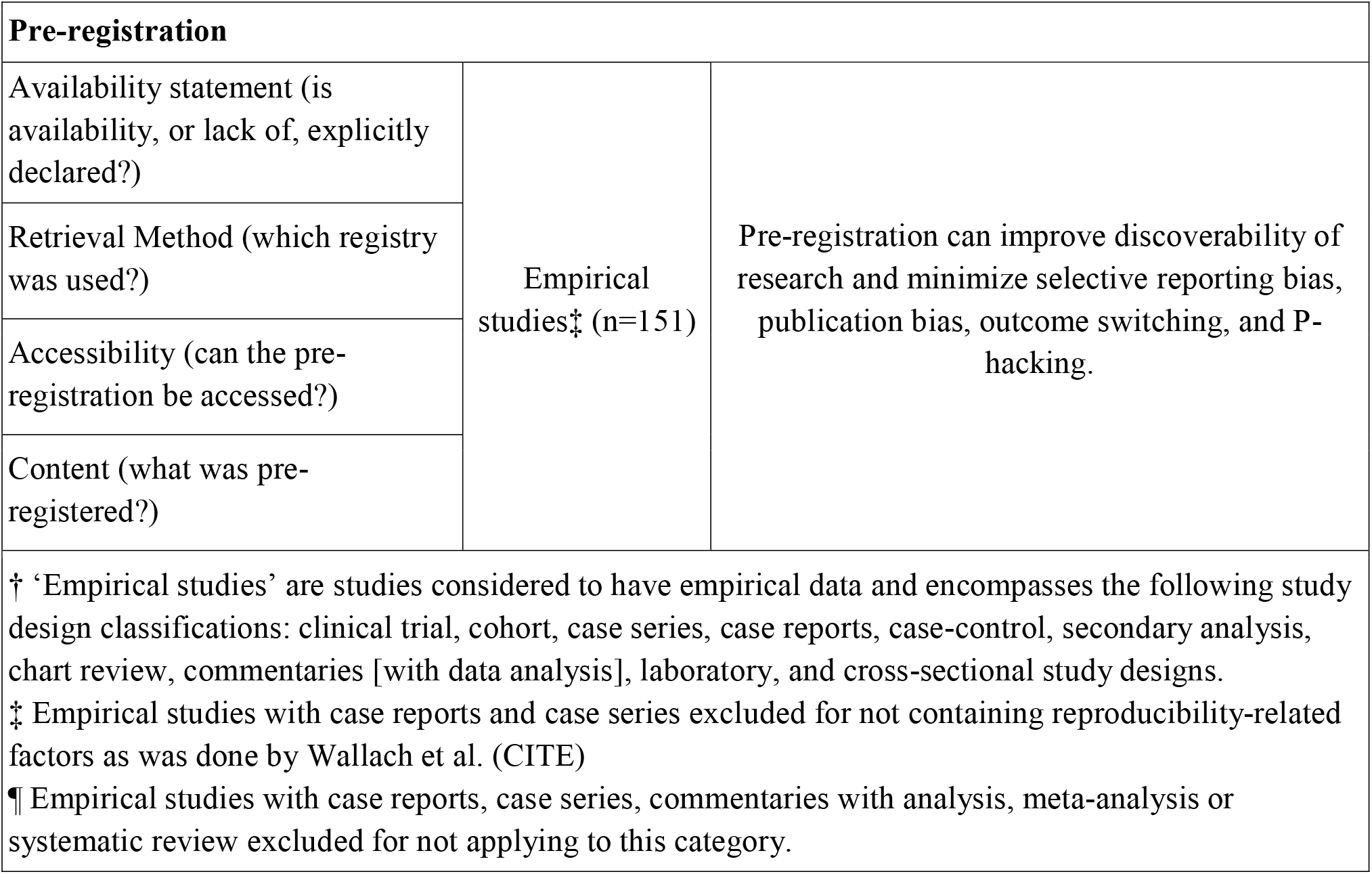
Measured variables. The variables measured for an individual article varied as it depended on the study design classification. Further details about extraction and coding is available here:https://osf.io/x24n3/

### Evaluation of Open Access Status

Open access was evaluated because many important components necessary for reproducibility are only available within the full text. Open access status for all 300 publications was evaluated by searching the publication’s title or DOI on the website Open Access Button (http://openaccessbutton.org). If publications could not be found on this platform, Google searches (https://www.google.com/) and PubMed searches (https://www.ncbi.nlm.nih.gov/pubmed/) were performed to verify open access status.

### Evaluation of Replication and Whether Publications Were Included in Research Synthesis

For empirical studies, we used Web of Science to determine whether the publication was replicated in other studies. For both empirical and nonempirical studies, we used Web of Science to determine the impact factor for each journal and whether the publication had been included in systematic reviews and/or meta-analyses. We performed these tasks by reviewing the title, abstract, and introduction of all publications in which the study was cited. This process was conducted in a duplicate, blinded fashion similar to our data extraction process.

### Data Analysis

Results are presented as frequencies and percentages along with 95% confidence intervals (95% CIs). All analyses were conducted using Microsoft Excel.

## RESULTS

Our initial search returned 26,498 records, from which 300 publications were randomly selected for analysis. Of the 300 randomly chosen publications, 286 were included and 14 were excluded either due to inaccessibility or because they had been retracted. Two hundred twenty-four publications had empirical data and 62 did not have empirical data. Furthermore, case reports and case series were excluded from publications with empirical data (n = 151) during select analysis (see Table 1). Figure 1 shows a flow diagram of included and excluded publications, and Table 2 presents the characteristics of the included publications.

**Table 2:** Study Characteristics for Included Publications.

| **Table 2: Study Characteristics for Included Publications** |  |  |
| --- | --- | --- |
| **Characteristic** |  | **Variables n (%)** |
| **Type of Study Included in Analysis (n=286)** | No empirical data | 62 (72.1) |
|  | Clinical trial | 43 (15.0) |
|  | Case series | 37 (12.9) |
|  | Case study | 36 (12.6) |
|  | Laboratory | 24 (8.4) |
|  | Cohort | 22 (7.7) |
|  | Chart Review | 17 (5.9) |
|  | Cross-sectional | 15 (5.2) |
|  | Survey | 10 (3.5) |
|  | Case control | 7 (2.4) |
|  | Meta-analysis | 5 (1.7) |
|  | Other | 4 (1.4) |
|  | Cost effect | 3 (1.0) |
|  | Commentary with analysis | 1 (0.3) |
| **Test Subjects (n=286)** | Humans | 203 (71.0) |
|  | Neither | 67 (23.4) |
|  | Animals | 15 (5.2) |
|  | Both | 1 (0.3) |
| **Country of journal publication (n=286)** | US | 167 (58.4) |
|  | UK | 65 (22.7) |
|  | Ireland | 23 (8.0) |
|  | Turkey | 6 (2.1) |
|  | Italy | 5 (1.7) |
|  | Brazil | 5 (1.7) |
|  | France | 3 (1.0) |
|  | Switzerland | 3 (1.0) |
|  | Unclear | 3 (1.0) |
|  | Other | 3 (1.0) |
|  | Netherlands | 2 (0.7) |
|  | Japan | 1 (0.3) |
| **Country of**<br>**corresponding**<br>**author (n=286)** | US | 100 (35.0) |
|  | Other | 66 (23.1) |
|  | UK | 20 (7.0) |
|  | China | 17 (5.9) |
|  | Japan | 16 (5.6) |
|  | Canada | 14 (4.9) |
|  | Turkey | 12 (4.2) |
|  | Germany | 11 (3.8) |
|  | Italy | 10 (3.5) |
|  | France | 9 (3.1) |
|  | South Korea | 8 (2.8) |
|  | Unclear | 3 (1.0) |
| **Replication**<br>**Studies (n=151)** | No statement | 151 (100) |
|  | Replication | 0 |
| **Cited by SR**<br>**and/or a Meta-**<br>**Analysis (n=224)** | No citations | 186 (83.0) |
|  | Not accessible† | 17 (7.6) |
|  | A single citation | 15 (6.7) |
|  | Two to five citations | 6 (2.7) |
| † Not accessible through web of knowledge to determine if publication was cited in a systematic review and/or a meta-analysis (www.webofknowledge.com) |  |  |

**Figure 1.**
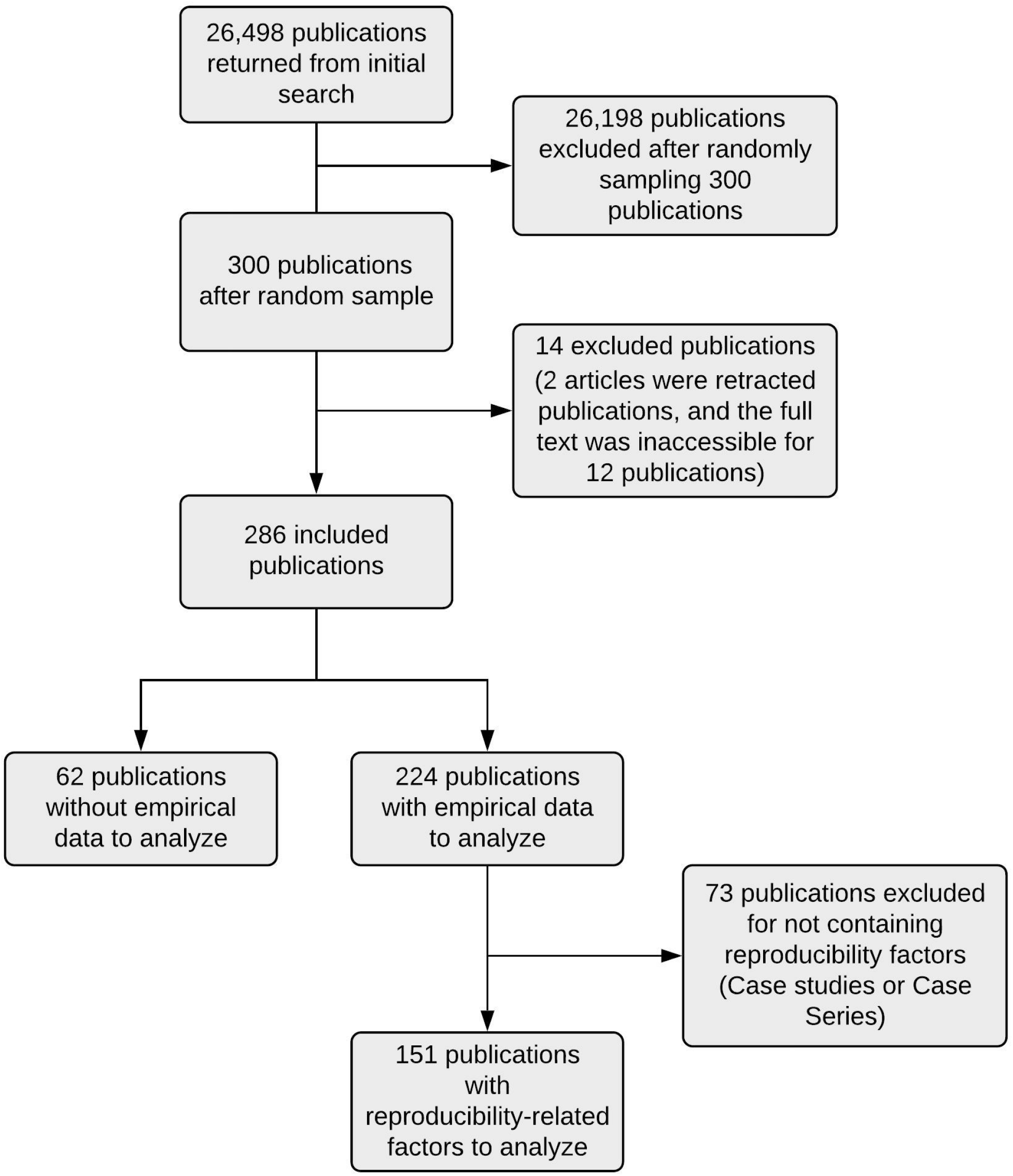
Flow diagram of included and excluded publications.

### Transparent and Reproducible Factors

Among the 300 publications analyzed, 67 (22.3% [95% CI, 17.6%-27%]) were available to the public using open Access Button, and 233 (77.7% [95% CI, 73-82.4]) were behind paywalls (Table 3). Of the 286 publications we could access, 59 (20.6% [95% CI, 16.1%-25.2%]) did not report a conflict of interest statement, 208 (72.7% [95% CI, 67.7%-77.7%]) reported no conflict of interest, and 19 (6.6% [95% CI,

**Table 3:**
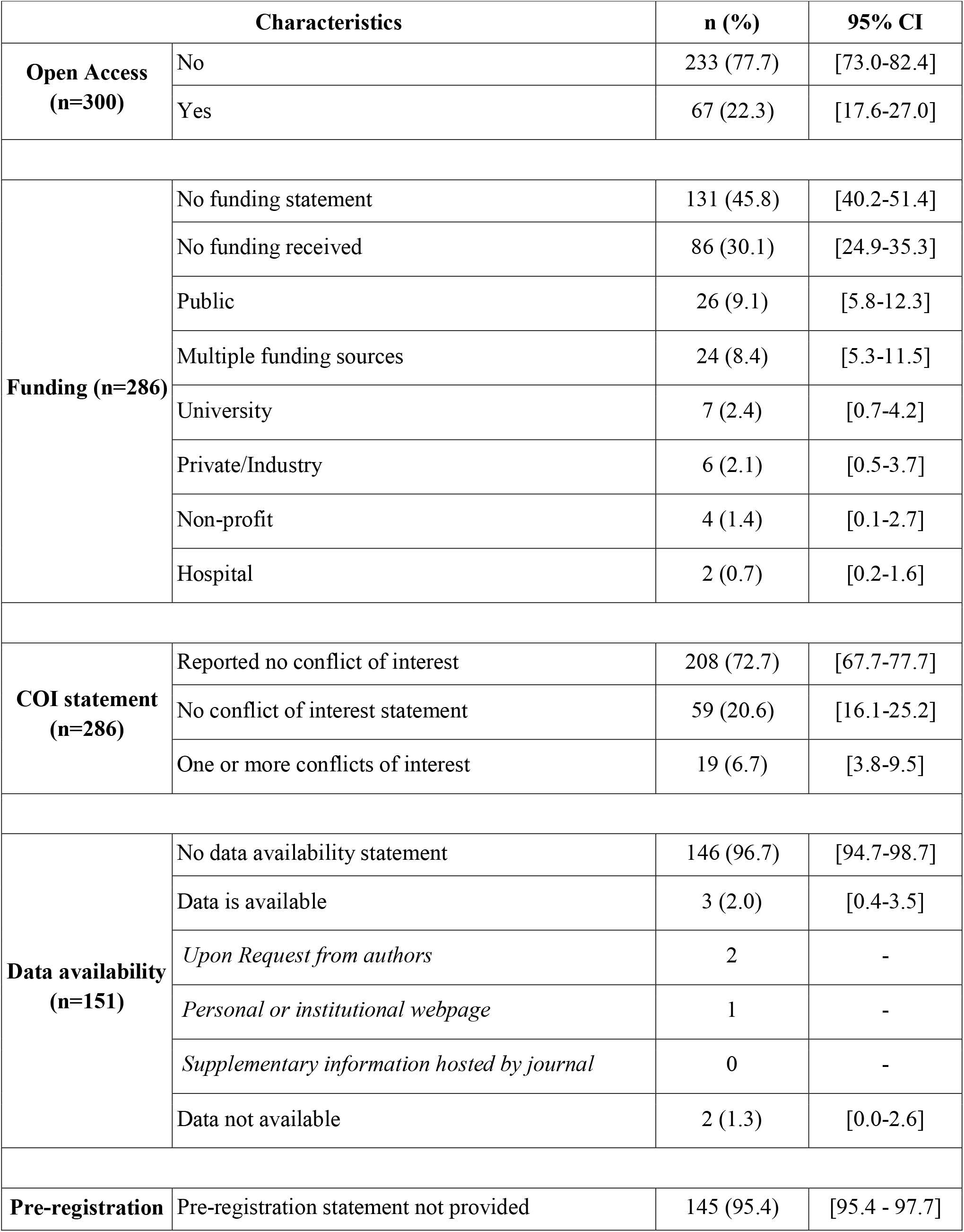
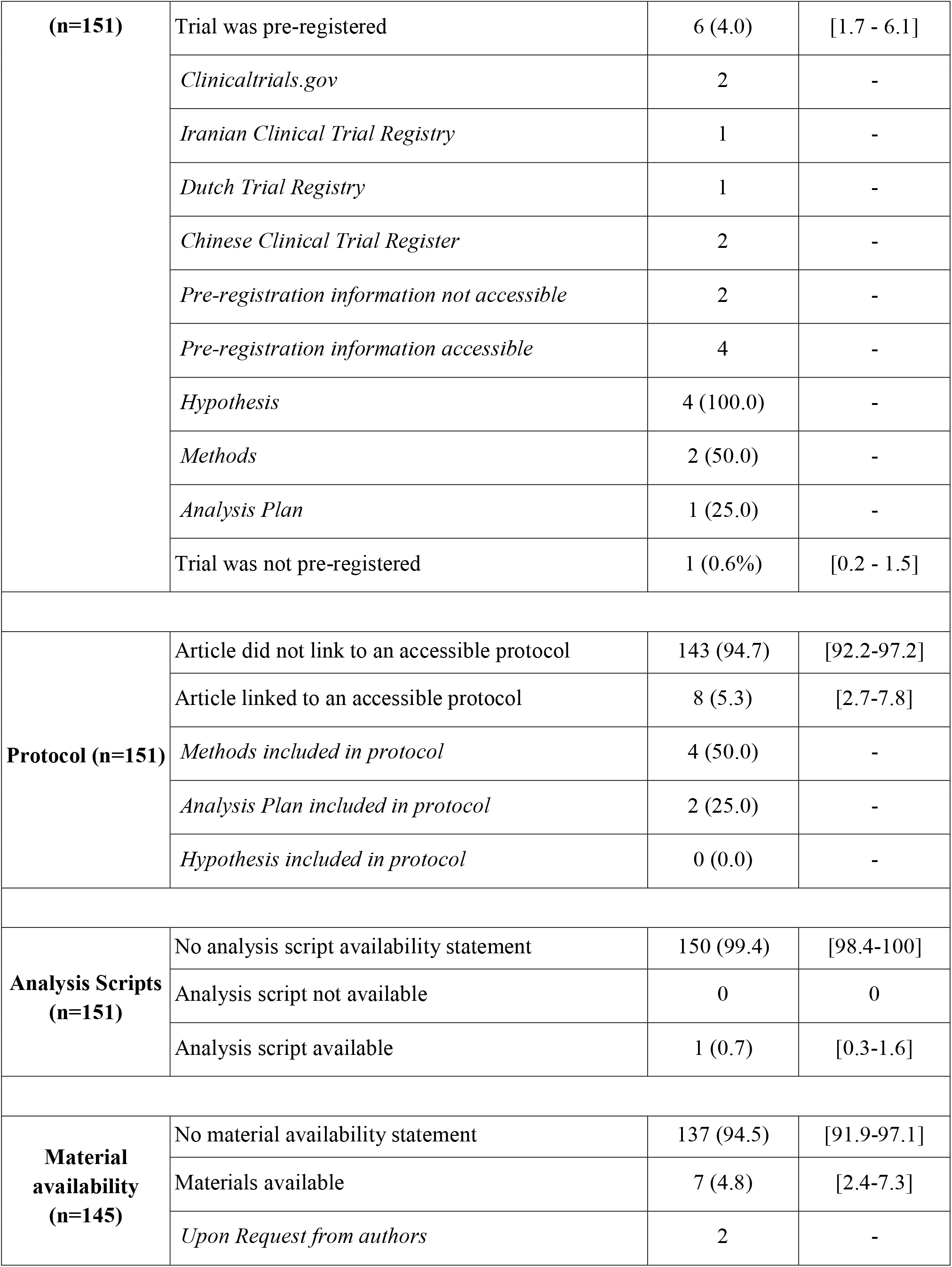
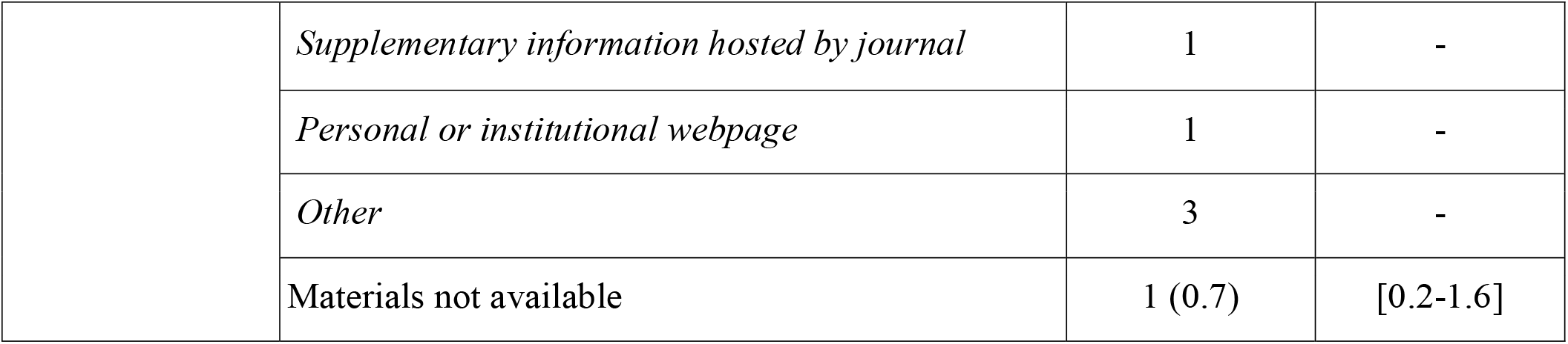
Factors Related to Reproducibility and Transparency.

3.8%-9.5%] reported 1 or more conflicts of interest. Further, 131 of the 286 publications (45.8% [95% CI,

40.2%-51.4%]) did not provide a funding statement, while 86 (30.1% [95% CI, 24.9%-35.3%]) stated they did not receive funding. Common sources of funding included public funding (26/286, 9.1% [95% CI, 5.8%-12.3%]), multiple funding sources (24/286, 8.4% [95% CI, 5.3%-11.5%]), and private/industry (6/286, 2.1% [95% CI, 0.5%-3.7%]). Of the 26 publications funded solely by public entities, only 9 (34.6%) were available as open access. Of the 224 publications that included empirical data, 21 (9.4%) were cited for analysis by a systematic review or meta-analysis.

Among 151 publications with empirical data (excluding case studies and case series), no publication had a clear statement claiming to replicate or be a replication of another study and only 6 (4.0% [95% CI, 1.7%-6.1%]) provided a preregistration statement. Similarly, only 3 (2.0% [95% CI, 0.4%-3.5%]) provided a data availability statement and 1 publication (0.7% [95% CI, 0.3%-1.6%]) provided an analysis script. Eight of the 151 publications (5.3% [95% CI, 2.7%-7.8%]) linked to an accessible research protocol. One hundred forty-five studies were eligible to be evaluated for material availability statements (6 were excluded for being commentaries [with analysis], systematic reviews, and/or meta-analyses). Of the 145 studies examined, 137 (94.5% [95% CI, 91.9%-97%]) did not contain a material availability statement. Complete analysis of transparent and reproducible factors is depicted in Table 3.

## Discussion

The majority of otolaryngology studies included in our analysis showed substandard procedures relating to reproducibility and transparency. Nearly all the studies lacked a data or material availability statement, did not link to an accessible protocol, and were not preregistered. In addition, most of the studies were not available through open access. Our findings align with previous literature in various fields of medicine.^15,22,23^ Many of these irreproducible studies lack specific methodology, resultant data, or access to materials needed to conduct the investigation.^9,24^ Irreproducibility carries an expensive price tag; for example, in their retrospective analysis, Freedman et al^25^ estimated that $28 billion has been spent on preclinical research that is irreproducible. Further, irreproducibility is a contributing factor in the growing public distrust in science.^22,25,26^ Complete transparency and reproducibility in research is likely unattainable, but advancement is possible and necessary.

One aspect of reproducible research is the availability of all the data and description of the materials necessary to conduct the experiment. While this study is the first to investigate data availability statements within otolaryngology, lack of data availability and sharing has already been shown to be an issue in other areas of medicine.^27^ We found that over 94% of analyzed studies did not provide a data or material availability statement. Similarly, an evaluation of 50 high impact factor journals in 2009 revealed that only 9% of studies supplied access to the full data online and 59% did not fully adhere to the data availability policy requirements.^28^ Such lack of access to raw data and access to materials hinders most efforts at reproducing a study. In response, some journals have begun requiring, or at least encouraging, the inclusion of data availability statements.^29^ These statements help facilitate replication of the study as well as reanalysis of its outcomes. Given the important role that data availability plays in reproducibility, we recommend that all journals adopt stricter policies on data availability as recently established by the International Committee of Medical Journal Editors (ICMJE) and studied in peer-reviewed journals such as *PLOS ONE* and *BMJ*.^27,29,30^ *PLOS ONE* has had a substantial increase over time in studies with data availability statements since enacting the policy.^29^ Otolaryngology journals, such as *Otolaryngology - Head and Neck Surgery* and *JAMA Otolaryngology - Head and Neck Surgery*,^31^ are listed as journals that follow ICMJE guidelines. We recommend all otolaryngology journals consider following suit.

Similar to data and material availability, a preregistration statement was not reported in over 95% of the studies that we analyzed. Preregistration is important because it helps guard against many threats to the integrity of scientific research.^32,33^ One specific threat that hinders research quality is selective outcome reporting bias. This bias occurs when some outcomes are reported and some are not, or when a predetermined secondary outcome is presented as the primary outcome.^34^ Reducing selective outcome reporting bias by preregistration is important in improving research quality overall, but also in improving research reproducibility. Studies become more difficult to reproduce when specific aims and outcomes are not defined a priori.

Another barrier to reproducibility is the lack of open access of published studies, which is especially concerning with regard to publications funded by public tax dollars. Over three-fourths of the studies analyzed in our investigation were not fully accessible, and of the 26 publicly funded studies, only 9 were available for full public access. If a study is financed by public tax dollars, a reasonable expectation is that the public should have unrestricted access to it in total.^35^ A general increase in open access research across otolaryngology would likely increase reproducibility within the field^36^ and may also improve the impact factors of otolaryngology journals, which tend to be relatively lower than journals in other areas of medicine.^36,37^ Our study corroborated the low impact factors, with a median 5-year impact factor of less than 2 being found for the analyzed otolaryngology journals. Multiple recent initiatives are currently attempting to address transparency, openness, and reproducibility in scientific literature.^21,33,38–41^

### Moving Forward

Given the reproducibility concerns within medical research, substantiated by our findings within otolaryngology, we support recommendations from the scientific literature to improve transparency and reproducibility-related factors (Figure 2). First, we recommend raising awareness of reproducibility issues and supporting behavioral changes through continued independent verification of data. Second, all peer-reviewed otolaryngology journals should consider requiring availability statements regarding materials, data sets, protocols, and analysis scripts (ie, factors related to reproducible research).^42–47^ Third, we propose prioritizing innovation of methods that facilitate data sharing compliance for researchers.^48^ Possible solutions include (1) using online repositories, such as Open Data for Science,^49^ Open Science Framework,^50^ or others (see http://www.re3data.org), (2) using OpenTrials,^51^ which is designed to share individual patient data from clinical trials in compliance with the Health Insurance Portability and Accountability Act of 1996, and (3) encouraging authors to prioritize preprint servers prior to submitting for publication. Lastly, academic institutions, peer-reviewed journals, and funding providers should consider incentivizing and rewarding investigators who fully disseminate materials, data sets, protocols, and analysis scripts by awarding specific levels or “badges” of reproducibility and transparency as laid out by the Center for Open Science.^52–54^ Badges are awarded for open data, open materials, and preregistration. This evidence-based incentive program is free to use for journals and organizations and has dramatically increased the rate of data sharing from 3% to 39% within psychological sciences.^55,56^ In our view, implementing these recommendations will support physicians in establishing credible and reliable research that ultimately governs clinical practice, as seen with the landmark Veterans Affairs study on chemoradiation versus surgery and radiation in laryngeal cancer.^57–59^ This study is a great example of reproducible research that has stood the test of time and improved patient care.

**Figure 2.**
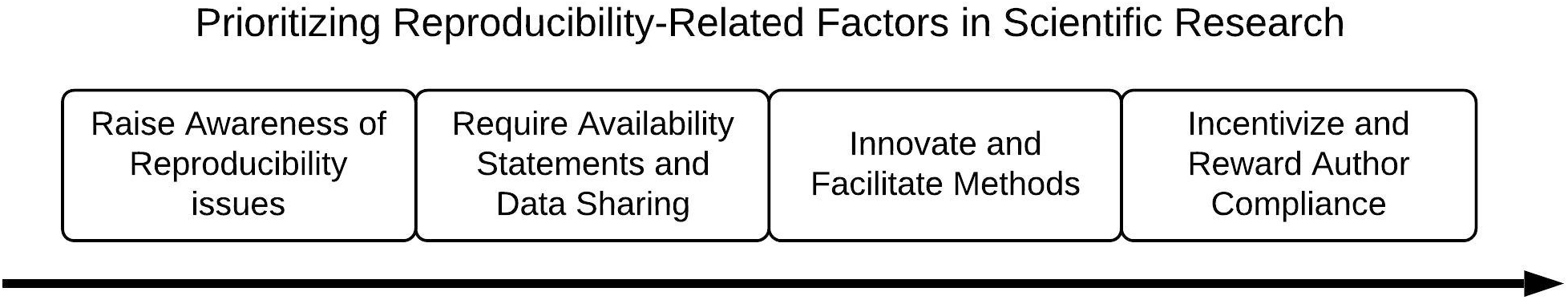
Recommendations for moving forward.

### Strengths and Limitations

Our study has both strengths and limitations. First, a lack of transparency may have been justified for some publications; however, justification was not made explicitly clear within the article. Second, we relied solely on published information for each article. We did not contact any authors because our objective was to measure the reported factors of transparency and reproducibility within published otolaryngology research. Third, the results of this cross-sectional review may not be generalizable to publications in other journals and time periods outside our search. Lastly, we are aware that research that does not employ reproducibility and transparency factors, as assessed in this study, may still add value to the otolaryngology community.

## Conclusions

Inadequate reproducibility practices exist in otolaryngology which hinders the practice of evidence-based medicine. Research should be conducted and presented in the same way one discusses a complicated case in consultation with a colleague—open and complete access to necessary information, with a genuine and unbiased presentation, to attain the best results.

## Data Availability

All protocols, materials, and raw data are available online.

https://osf.io/x24n3/

